# Feasibility of Longitudinal Relaxation Rate Mapping with Non-Cartesian Sampling and Compressed Sensing on a 1.5T MR-Linac

**DOI:** 10.1101/2025.07.28.25332213

**Authors:** Lucas McCullum, Michael J. van Rijssel, Ken-Pin Hwang, Yao Ding, Chad Tang, Comron Hassanzadeh, Jinzhong Yang, Peter A. Balter, Jihong Wang, Clifton D. Fuller, Ergys D. Subashi

## Abstract

**Background:** Quantitative mapping of the longitudinal relaxation rate (R1=1/T1) is a major building block for several multiparametric MRI protocols intended for adaptive radiation therapy planning. The implementation of these protocols is challenging in anatomical sites that experience large physiological motion.

**Purpose:** To implement and validate a motion-resolved quantitative T1 mapping method on a 1.5T MR-Linac that combines non-Cartesian k-space sampling trajectories with compressed sensing (CS) reconstruction techniques.

**Methods:** Four 3D non-Cartesian k-space trajectories were evaluated: radial and stack-of-stars sampling using half- and full-spoke coverage. A variable flip angle acquisition was performed using the spoiled gradient-echo sequence, and T1 mapping was validated using two standard phantoms. Gradient delay timing was optimized empirically to minimize trajectory-induced artifacts. Eight compressed sensing reconstruction strategies were tested using spatial and spatiotemporal regularization operators. Reconstructions were evaluated across multiple implementation parameters and ranked based on spatial resolution, bias, and variability. In vivo studies included one healthy volunteer and one patient undergoing radiotherapy to a target in the kidney. Motion-resolved imaging was performed using respiratory self-gating and phase-sorted reconstruction.

**Results:** All non-Cartesian trajectories demonstrated high repeatability and low longitudinal bias in phantom studies, with coefficients of variation below 3.3%. Radial half-spoke sampling achieved the shortest scan times and highest agreement with Cartesian benchmarks. Reconstruction methods incorporating spatiotemporal regularization maintained spatial resolution and quantitative accuracy across undersampling factors up to 20-fold. In human subjects, non-Cartesian T1 mapping provided improved accuracy and reduced variability in mobile abdominal tissues compared to Cartesian acquisitions, particularly in the kidney cortex and medulla, where motion artifacts led to overestimation and higher variance in the reference method.

**Conclusions:** T1 mapping using non-Cartesian trajectories and compressed sensing reconstruction is feasible on a 1.5T MR-Linac. The proposed approach enables accurate, motion-resolved quantitative imaging within clinically practical acquisition times. These results support integration of quantitative T1 mapping into adaptive MR-guided radiotherapy workflows and establish a foundation for future development of multiparametric imaging and response-adaptive treatment strategies.

## 1. Introduction

The integrated Elekta Unity MR-Linac (Elekta AB; Stockholm, Sweden) provides a novel platform for precision radiotherapy, combining a diagnostic 1.5T magnetic resonance imaging (MRI) scanner with a 7 MV linear accelerator (Linac)^1^. This system enhances the accuracy in the delineation of targets and organs-at-risk (OARs) due to its high spatiotemporal resolution and superior soft tissue contrast compared to conventional CT-based methods. MR-guided adaptive radiotherapy (MRgART) as a field, of which the MR-Linac belongs, further supports several online adaptive strategies, optimizing radiation dose delivery to the target while minimizing risks to adjacent normal tissues. The efficiency of online MRgART is anticipated to further benefit from automation techniques for treatment planning, staging, and outcome modeling^2^.

The versatility of several MRI pulse sequences and their corresponding contrast mechanisms may significantly improve radiotherapy outcomes by enabling noninvasive mapping of functional^3^, physiologic^4^, and metabolic biomarkers^5^. Multiparametric MRI protocols complement and extend current diagnostic tests for detecting and characterizing malignancies with strong correlations to histological validation^6^. Furthermore, recent research emphasizes the importance of biologically informed adaptive planning, integrating spatiotemporal variations in tumor biology into MRgART workflows. Specifically, changes in the longitudinal relaxation rate (R1 = 1/T1) have been used to predict outcomes in glioblastoma patients^7^ and evaluate radiation response in the heart^8^. Additional work is beginning to focus on how changes in the longitudinal relaxation rate affect normal tissue as well^9^.

Longitudinal relaxation rate mapping is a major building block for several multiparametric MRI protocols^10–12^. However, the implementation of these protocols is challenging in anatomical sites that experience large physiological and respiratory motion leading to significant challenges in radiotherapy, particularly for targets in the thorax and abdomen^13^. MRI pulse sequences relying on non-Cartesian sampling, specifically radial (projection-encoding) methods, offer intrinsic advantages in clinical imaging. Radial sampling provides inherently volumetric data, reduces sensitivity to flow artifacts, and enables shorter echo times compared to Cartesian techniques. When the Nyquist criterion is satisfied across the entire domain in k-space, these sampling functions significantly enhance image quality for regions with high amounts of motion with minimal streaking artifacts^14^. Additionally, radial k-space trajectories inherently encode the respiratory motion signal, eliminating the need for external tracking devices, a method known as self-gating^15^. The inherent redundancy in projection encoding facilitates the implementation of a range of advanced reconstruction methods derived from view-sharing^16^, compressed sensing^17^, and machine learning algorithms^18^ enabling reduced acquisitions times at higher spatial and temporal resolution. Several combinations of these methods have been employed such as StarVIBE^19^, GRASP^20^, and XD-GRASP^21^, among many others.

The current problem of implementing non-Cartesian T1 mapping on the MR-Linac is the limited time available for acquisition and limited existing literature comprehensively investigating the optimal methodologies in a time-constrained setting. Therefore, in this study, we investigate four different non-Cartesian k-space trajectories and eight different offline compressed sensing algorithms to determine the optimal combinations that provide the highest quantitative T1 mapping accuracy at the highest level of undersampling with minimal imaging artifacts.

## 2. Materials and Methods

### 2.1. Acquisition and Reconstruction Strategy

A robust implementation of longitudinal relaxation rate mapping using non-Cartesian MRI relies on the ability to (1) estimate a physiologic signal that allows for matching motion states and for retrospective correction of motion-induced artifacts, (2) design an acquisition trajectory to ensure sparse and quasi-uniform k-space sampling for each motion state, and (3) identify a compressed-sensing reconstruction method with a regularization term that optimizes image quality in the spatial and parameter domain. In the context of simulation and treatment planning for targets in the thorax and abdomen, three-dimensional (3D) radial trajectories with a randomized distribution of polar and azimuthal angles can be designed to sample each respiratory phase while ensuring full k-space coverage with minimal overlap between sampling points. The sampling functions enable randomized acquisitions that provide efficient k-space coverage across all respiratory phases. These functions can also sample the center of k-space at each repetition time, allowing for the measurement of a motion-correlated signal that derives from the average magnetization in the excited field-of-view^22^.

### 2.2. Validation of Clinical Methods

The initial feasibility of longitudinal relaxation rate mapping using clinical non-Cartesian pulse sequences and reconstruction was evaluated at two institutions. Phantom experiments were performed on the integrated 1.5T MR-Linac (Unity; Elekta AB; Stockholm, Sweden) running Philips software version R5.7.1.2. Quantitative T1 mapping was implemented using the variable flip angle (VFA) method^23,24^ with a radiofrequency (RF) spoiled (T1-enhanced) gradient echo sequence (SPGR). The optimal flip angles were determined using DESPOT1 theory^25^ by taking the mean across T1 values in the range of 250 – 2000 ms in line with typical clinical values in the abdomen^26^. Additionally, the flip angle used during the preparation phases (Larmor frequency determination, power optimization, DC correction, coil tuning and matching) was determined by calculating the respective mean Ernst angle^27^ across the desired range of T1 values. Two different 3D non-Cartesian k-space sampling strategies were investigated: pseudo-randomized radial “kooshball” and stack-of-stars. Further, each non-Cartesian k-space sampling strategy was explored using both a half-spoke (sampling begins at the center of k-space) and full-spoke (sampling begins at the edge of k-space) trajectory as shown in **Figure 1**.

**Figure 1:**
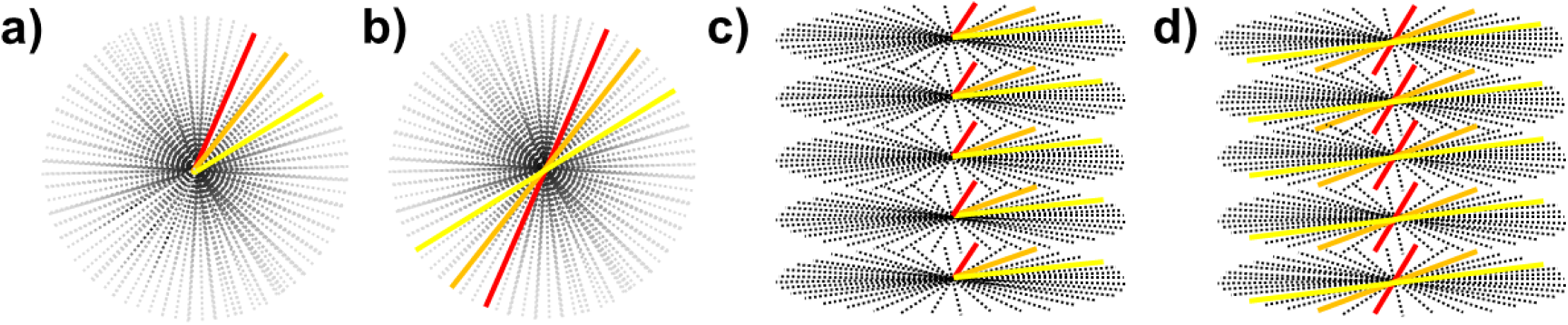
The 3D k-space sampling trajectories investigated in this study: (a) half-spoke radial, (b) full-spoke radial, (c) half-spoke stack-of-stars, and d) full-spoke stack-of-stars. Note, the colored lines represent a single readout where half-spoke acquisitions begin at the center of k-space and full-spoke acquisitions span the range from -k_max_ to +k_max_.

For performance benchmarking and comparison within anatomical regions with physiological motion, a Cartesian k-space acquisition was included. The total acquisition time was maintained within 5 minutes to accommodate typical clinical scan time requirements and workflow optimization for adaptive radiotherapy. Scan times well below this target were achievable for the full-spoke acquisitions. For each non-Cartesian sequence, a TR of 10 ms was maintained and the minimally achievable TE was selected. Relevant imaging parameters are summarized in **Table 1**. The re-gridding algorithm with Sum-of-Squares reconstruction^28^ was used as implemented by default in the MR-Linac.

**Table 1:**
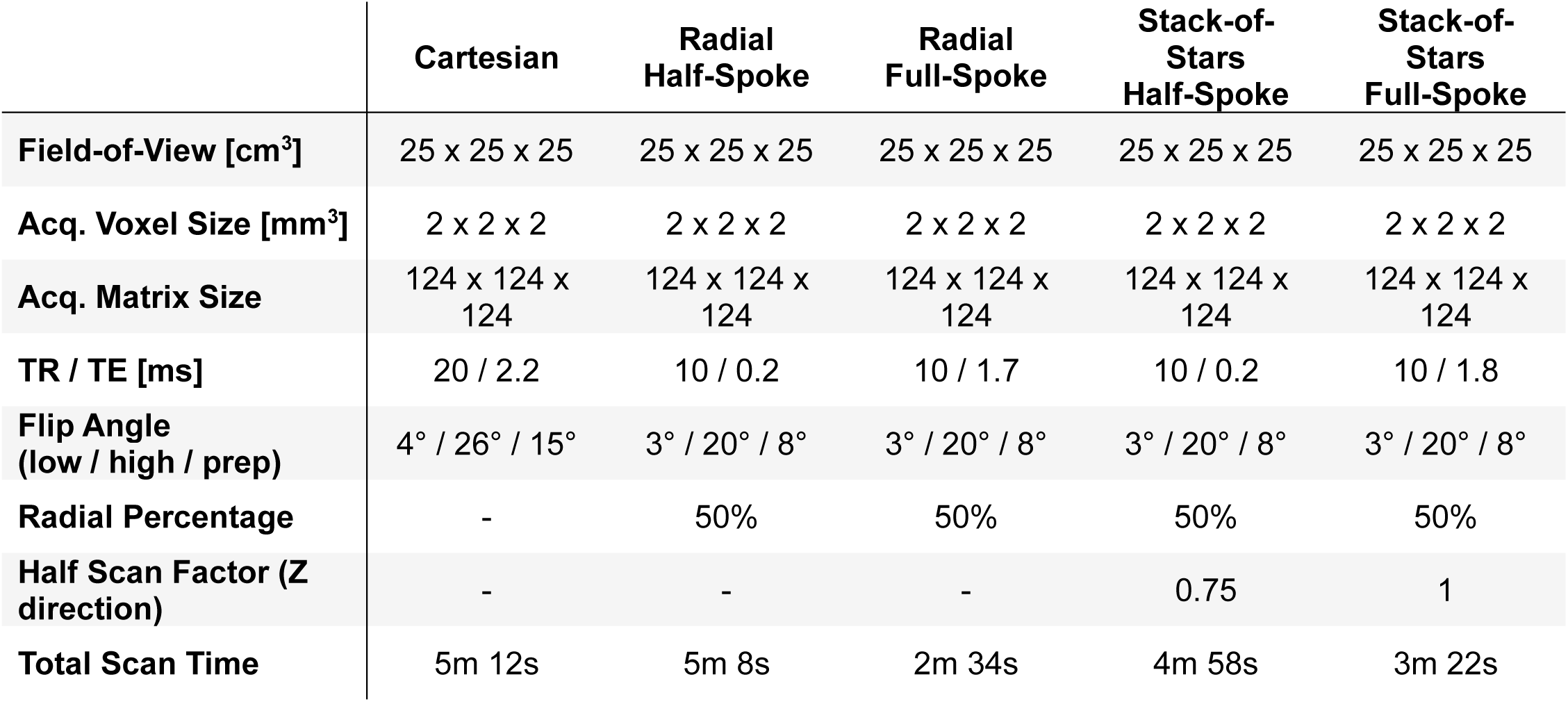
MRI sequence parameters used for T1 mapping with the VFA method. A consistent acquisition geometry was maintained to allow easier comparison between the different sampling methods and avoid additional confounders. A constant TR and minimally achievable TE was maintained for each non-Cartesian acquisition. To keep all measurements within ∼5 minutes, a 50% radial sampling percentage was selected, and the half-spoke stack-of-stars trajectory required an additional 75% half scan factor in the z-direction. The total scan time including the preparation and both flip angles is shown in the last line.

Two phantoms were used for quantifying the bias, repeatability, and reproducibility of the T1 mapping methods. In one institution, the CaliberMRI “ISMRM/NIST” Premium System Phantom (Model 130; CaliberMRI; Boulder, CO) was used as a reference for NIST-traceable T1 values^29^. This phantom includes 28 reference vials with nominal T1 values in the range 22 – 1898 ms at 20°C and 1.5T. Scanner temperature was verified using a thermometer located next to the bore and maintained in the range 19 – 22°C. Repeat scans were acquired over five days to evaluate longitudinal bias and reproducibility. The other institution used the MagIQ Eurospin II TO5 Phantom^30^ (Leeds Test Objects; Boroughbridge, UK) which includes 12 vials suitable for nominal T1 values in the range 195 – 1599 ms at 20°C and 1.5T. Bias was assessed with respect to the T1 values measured with the benchmark method, while reproducibility was assessed using the longitudinal coefficient of variation (CoV) for the repeat scans. Spearman’s test was used to evaluate the correlation of bias with T1 as a measure of R1-dependence on accuracy of acquisition method.

### 2.3. Gradient Delay Optimization

Non-Cartesian k-space acquisitions are susceptible to minor deviations in the sampling trajectory arising from timing errors in the gradient system electronics^31^. Minimizing the discrepancy in timing parameters between expected and actual gradient ramping also minimizes trajectory-mismatch artifacts which typically appear as asymmetric ghosting artifacts at the edge of the image. This discrepancy is dependent on the gradient system and can be determined empirically by varying the gradient delay timing parameter. Ghosting artifacts due to gradient delays were estimated by generating 360 radially spaced (1° separation) line profiles at the edge of the ISMRM/NIST phantom. The edge ghosting artifact was quantified by comparing the mean signal intensity just inside the phantom edge to just outside the phantom edge. The ratio between the mean signal intensity outside the phantom edge to mean signal intensity inside was used as an estimate of the magnitude of ghosting artifact. This ratio was evaluated for gradient trajectory delays in the range 0 – 5 μs. The optimal delay was determined based on the minimum ratio of signal intensities outside and inside the phantom.

### 2.4. Offline Compressed Sensing Reconstruction

In this work, we adopt the formalism of parallel imaging with compressed sensing (PICS) as provided in the implementation of the Berkley Advanced Reconstruction Toolbox^32–35^ (BART):

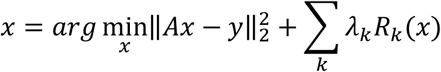

where *x* = unknown image, *y* = measured k-space data, *A* = imaging acquisition model, *λ* = regularization parameter, *R* = regularization operator, and *k* = number of regularization operators^36^. This formalism allows for the combination of several regularization operators along the spatial, temporal, or spatiotemporal domain. One of the primary motivations for implementing this reconstruction approach is its historical evaluation and acceptance for clinical use in conventional MRI simulators. Nevertheless, while compressed sensing reconstruction with various regularization operators has been thoroughly studied for anatomical imaging, its use in the MR-Linac for quantitative imaging remains work in progress. We hypothesize that an optimized implementation of PICS in the MR-Linac allows for a large degree of undersampling in non-Cartesian sequences used for longitudinal relaxation rate mapping. However, analytical methods for identifying the optimal regularization operator and its respective regularization parameters are possible only for a limited number of regularizers. In practice, the optimal operators are found through an iterative, empirical, and calibration process focusing on the optimization of image metrics in the domain of interest. In our study, the undersampling factor was varied by a factor of 5, 10, 15, 20 and the optimal regularization parameter was identified based on an L-curve search^37^. The search was initially focused on maximizing resolution in the spatial domain, followed by a search for minimizing bias and CoV in the parameter (T1) domain. The regularization operators studied in this work are listed in **Table 2**, the regularization parameter was varied in the range 10^−4^ – 10^−1^, and the number of iterations was varied between 25, 50, 75, and 100. In each acquisition, coil sensitivity profiles were computed using ESPIRiT by combining all available k-space spokes^38^.

**Table 2:** A summary of compressed sensing algorithms used for offline reconstruction. In total, six unique regularization transforms (Fourier, Image, Wavelet, Local Casorati, Finite Differences, and Two-Level Finite Differences), three unique penalty functions (L1 Norm, L2 Norm, and Nuclear Norm), and two unique transform domains (Space and combined Space/Time) were tested in various combinations. Corresponding shorthand abbreviations are shown on the left column.

| Symbol | Regularization Transform | Penalty Function | Transform Domain |
| --- | --- | --- | --- |
| L1F<sub>s</sub> | Fourier | L1 Norm | Space |
| L1<sub>s</sub> | Image | L1 Norm | Space |
| L2<sub>s</sub> | Image | L2 Norm | Space |
| L1W<sub>s</sub> | Wavelet | L1 Norm | Space |
| L1W<sub>ST</sub> | Wavelet | L1 Norm | Space + Time/Contrast |
| LLR<sub>ST</sub> | Local Casorati | Nuclear Norm | Space + Time/Contrast |
| TV<sub>s</sub> | Finite Differences | L1 Norm | Space |
| TGV<sub>s</sub> | Two-Level Finite Differences | L1 Norm | Space |

As in the experiments on gradient delay optimization, we generated 360 radially spaced (1° separation) line profiles at the edge the ISMRM/NIST phantom image. The full-width half-maximum (FWHM) of the gradient of the line profile was used as a metric for spatial resolution. Bias was assessed with respect to T1 values measured with the fully sampled reconstruction, and uncertainty was assessed using the CoV across the range of T1 values considered in this work.

### 2.5. In Vivo Imaging

The feasibility of the sampling and reconstruction method were evaluated in one healthy volunteer and one patient undergoing radiation therapy for a tumor in the left kidney. Imaging parameters are listed in **Table 1**. To accommodate patient size, the acquired FOV and sampling matrix were changed to 35 x 35 x 35 cm^3^ and 116 x 166 x 116. The change in acquisition sampling matrix allowed for a scan time of ∼2.2 minutes per flip angle. All images were reconstructed at 1 mm^3^ isotropic resolution. In vivo experiments used the 3D radial sampling function with the half-spoke trajectory and optimal gradient timing delay. The respiratory signal was identified by principal component analysis (PCA) of the signal extracted from the center of k-space from all eight receive RF-channels. PCA was performed along the channel direction, identifying the receive signal with the strongest representation of the respiratory signal. Thereafter, the respiratory signal was phase or amplitude-sorted into a user selected number of breathing phases. The re-gridding algorithm with Sum-of-Squares reconstruction was used as reference, and offline PICS was implemented with the optimized regularization operator and parameters identified in the search process in the phantom experiments. A body mask was defined based on thresholding the low flip angle acquisition and the parameter maps were filtered to remove noise outside the patient. Research subjects were consented to an internal imaging protocol (PA15-0418) as approved by the institutional review board at The University of Texas MD Anderson Cancer Center.

### 2.6. Image Analysis

For the phantom analysis, a circular region-of-interest (ROI) with a 1 cm diameter was created at the center of each vial using 3D Slicer^39^. The T1 maps were reconstructed offline by following the DESPOT1 formalism. Bias and reproducibility was assessed using both the ISMRM/NIST and Eurospin phantom and quantified using the percent difference from benchmark values and coefficient of variation, respectively. Longitudinal repeatability was assessed in the ISMRM/NIST phantom by acquiring repeated scans over five days and quantified using the median bias and coefficient of variation across repeat scans. Spearman’s rank correlation coefficient^40^ was used to evaluate the correlation of median bias across five days with measured T1 within each respective phantom vial. The Lin’s Concordance Correlation Coefficient^41^ (LCCC) was calculated as a measure of agreement. The analysis was constrained in vials with T1 relaxation times in the range of 250 – 2000 ms and T2 relaxation times in the range of 0 – 200 ms^26^.

## 3. Results

### 3.1. Validation of Clinical Methods

The ISMRM/NIST phantom contains six vials with T1 values in the range 250 – 2000 ms and T2 values in the range 0 – 200 ms, while the Eurospin phantom contains 11 vials. **Figure 2a-b** shows representative T1-maps from each phantom. The T1-maps in this example are calculated from datasets acquired with half-spoke and full-spoke trajectories. **Figure 2c** plots the relationship between mean T1 measured with the reference Cartesian acquisition and the non-Cartesian sampling method.

**Figure 2:**
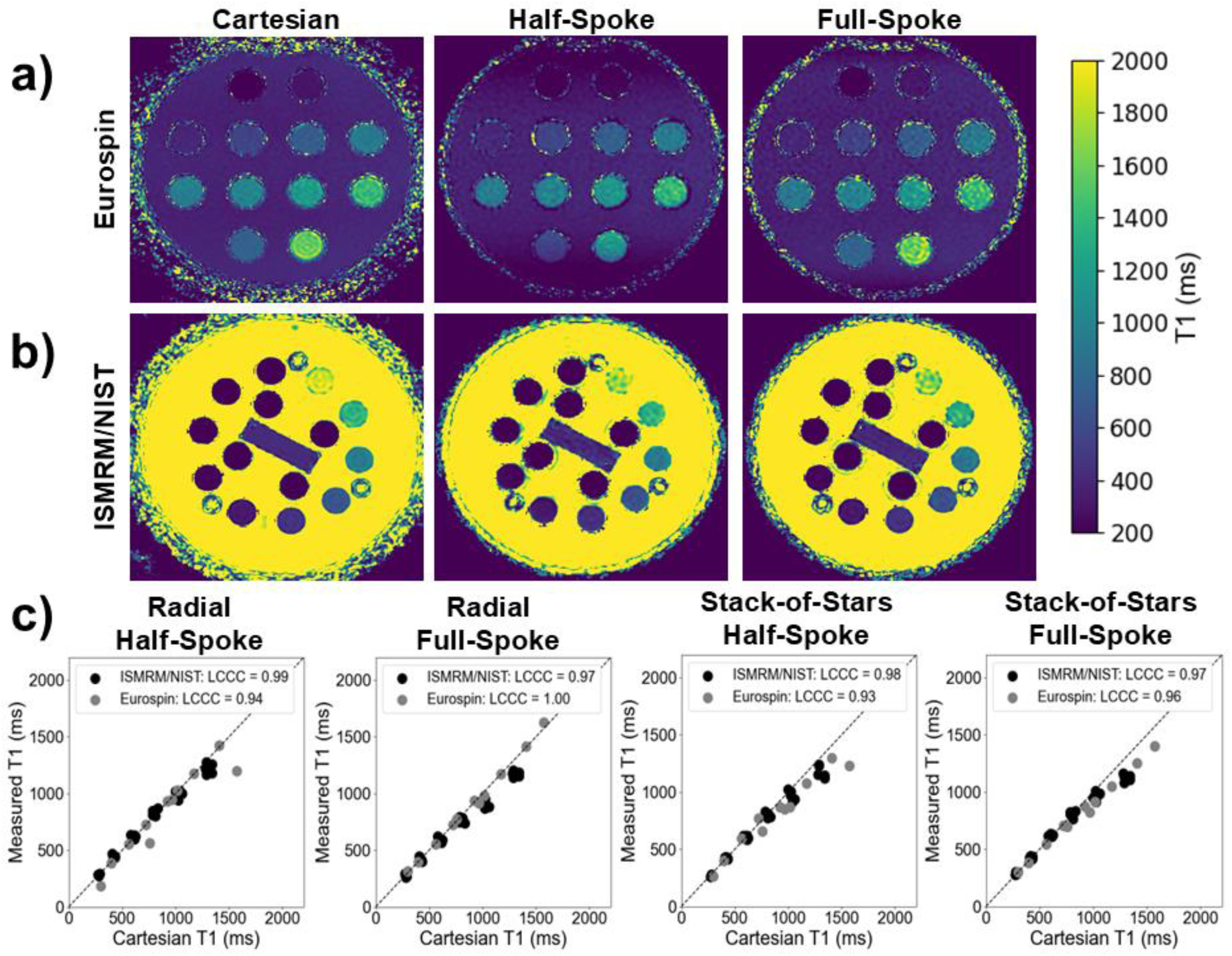
(a) Representative T1 maps in the Eurospin phantom and (b) ISMRM/NIST phantom for the Cartesian (left), half-spoke radial (middle), and full-spoke radial (right) sampling. (c) Measured vs. Cartesian T1 values in both the ISMRM/NIST and Eurospin phantom across each sequence and their respective LCCC accuracy quantification metrics.

The analysis of the data for determining the optimal gradient delay revealed that edge ghosting artifacts were minimized at a timing delay of 3 µsec for both 3D radial and stack-of-stars sampling methods. **Figure S1** in Supplementary Materials shows an example of the edge ghosting artifact for an acquisition with no gradient delay timing and demonstrates the change in line profiles as a function of increase in the value of the gradient delay. Based on this data, all imaging experiments in phantoms and in vivo were performed using a gradient trajectory delay of 3 μs.

**Table 3** lists the bias and reproducibility metrics for comparisons with respect to the Cartesian benchmark and longitudinally. All sampling methods have negligible longitudinal bias with a maximum coefficient of variation ∼ 3.3%. Additionally, the correlation between bias and measured T1 is not statistically significant for the half-spoke measurements while significant for the full-spoke. The full-spoke acquisitions demonstrated a negative correlation with bias as the T1 values increased. Although a high concordance coefficient is observed, the stack-of-stars sequences exhibited an underestimation of T1 at the largest reference values measured with the Cartesian sequence. A similar trend was noted in the full-spoke radial sequence, though less severe. This effect is due to insufficient spoiling and signal recovery as can be seen in the ISMRM/NIST phantom with vials of high T1 and T2 values, shown in **Figure S2** in the **Supplementary Materials**.

**Table 3:** Comparison of bias and reproducibility metrics with respect to Cartesian benchmark. Longitudinal repeatability was assessed in the ISMRM/NIST phantom by acquiring repeated scans over five days and quantified using the median bias and coefficient of variation across repeat scans. Spearman’s rank correlation coefficient was used to evaluate the correlation of median bias with measured T1.

|  | Radial<br>Half-Spoke | Radial<br>Full-Spoke | Stack-of-Stars<br>Half-Spoke | Stack-of-Stars<br>Full-Spoke |
| --- | --- | --- | --- | --- |
| Median Bias / CoV vs. Cartesian (%) | 0.8 / 7.4 | -6.9 / 8.2 | -3.7 / 8.0 | -0.6 / 6.9 |
| Longitudinal Bias / CoV (%) | 0.1 / 2.8 | -0.03 / 3.3 | 0.6 / 2.3 | 0.3 / 2.5 |
| Spearman’s Test for Bias vs. T1 (p-value) | 0.21 | 0.005 | 0.40 | 0.04 |

### 3.2. Offline Compressed Sensing Reconstruction

**Figure 3** presents a comparative example of the variation in image quality in the ISMRM/NIST phantom as a function of undersampling factor and reconstruction method. **Figure 3a** shows the phantom image reconstructed with no undersampling, **Figure 3b-d** display images reconstructed with the reference method, while **Figure 3e-g** display results with compressed sensing reconstruction with L1-wavelet regularization in the spatial and temporal (i.e. contrast) domain. The undersampling factor in these examples is x10 and x20. The calculated T1 maps are shown in the range 200 – 4000 ms to further demonstrate the effect of undersampling and reconstruction on bias and signal-to-noise.

**Figure 3:**
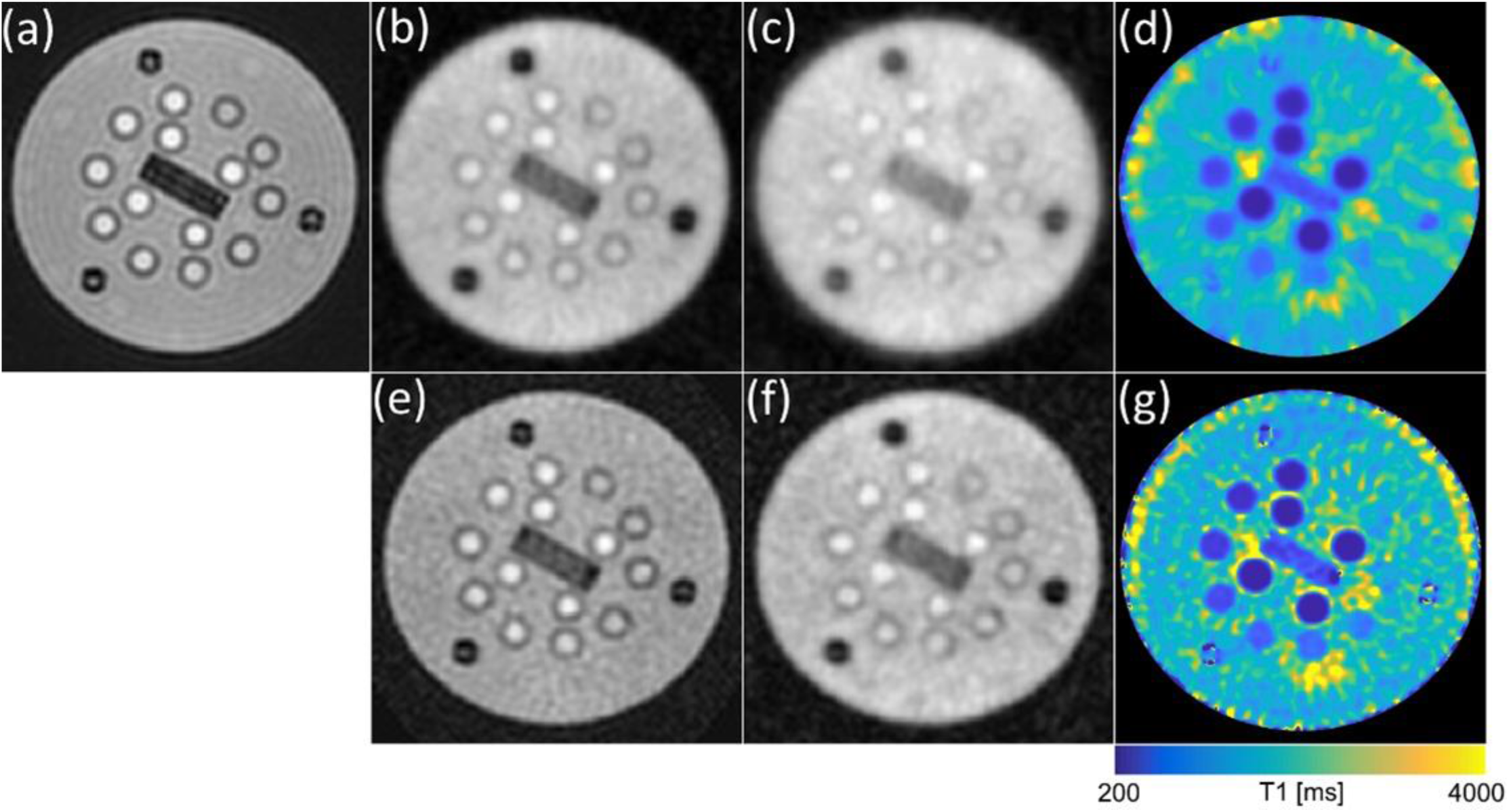
Representative dataset from acquisition with 3D half-spoke radial sampling reconstructed with (a-d) sum-of-squares re-gridding and (e-g) compressed sensing with L1W_ST_. The figure panels represent (a) fully sampled, (b,e) 10x undersampling, (c,f) 20x undersampling, (d,g) calculated T1 map with 20x undersampling.

The effect of undersampling and reconstruction method on spatial resolution was quantified using the FWHM of line profiles at the edge of the ISMRM/NIST phantom. The initial search for optimal regularization operator and parameter was focused on maximizing spatial resolution. For a given undersampling factor, reconstruction methods were ranked based on highest achieved spatial resolution, followed by bias and CoV. **Figure 4** plots the median FWHM vs rank as a function of undersampling and regularization operator. The highest rank method achieved the smallest median FWHM. When two methods achieved equal values of FWHM then they were ranked based on bias and CoV. As the undersampling factor is increased, there is a clear distinction in robustness between two groups of regularization operators. L1-regularization in the wavelet/Fourier domain and LLR maintain similar spatial resolution across undersampling factors. Regularization in the image domain is not able to recover the decrease in spatial resolution at high undersampling factors.

**Figure 4:**
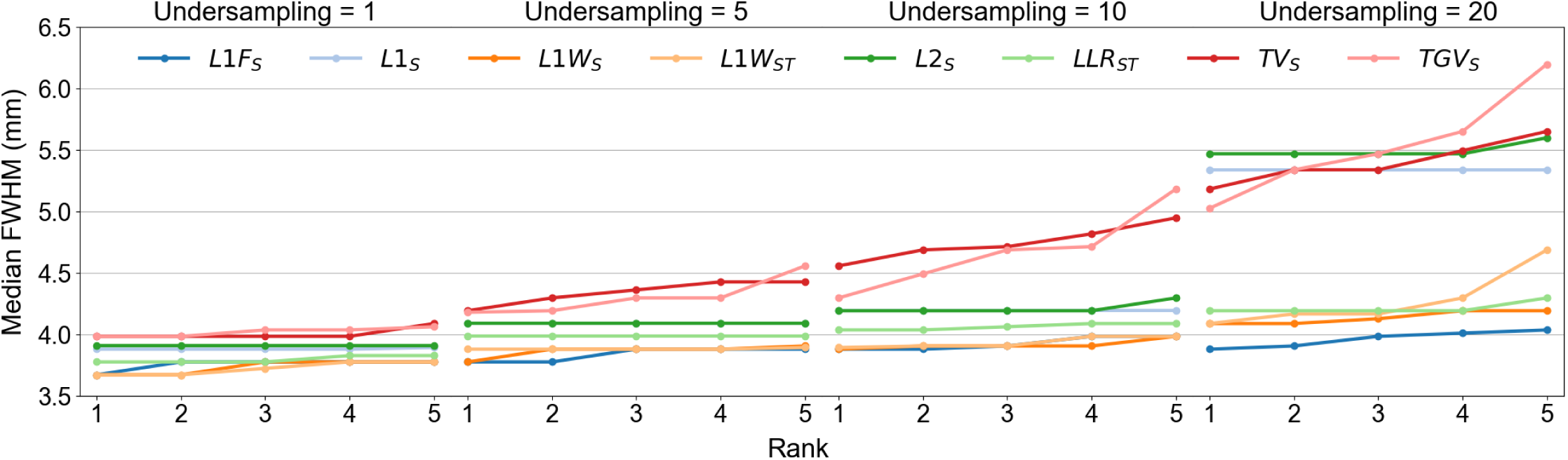
Spatial resolution as a function of undersampling and reconstruction method. The highest rank method achieved the smallest median FWHM. When two methods achieved equal values of FWHM then they were ranked based on bias and CoV. Notice a separation in robustness between two groups of regularization operators as the undersampling factor is increased.

The reconstruction methods using L1F_S_, L1W_S_, L1W_ST_, and LLR_ST_ were robust to changes in the undersampling factor and were further evaluated based on bias and CoV in calculated T1 maps. **Figure 5** shows bias with respect to T1 maps calculated from fully sampled datasets and CoV as a function of regularization parameter across all undersampling factors. This example presents results for T1 maps calculated from images reconstructed using joint L1-wavelet regularization in the spatial and contrast domain (i.e. L1W_ST_). For this regularization operator, uncertainty across T1 maps is minimized at regularization parameters in the range 10^−3^ – 10^−2^ irrespective of undersampling factor. Furthermore, bias is constant once the regularization parameter is less than 10^−2^. This behavior is also generally observed in the other regularization parameters, except for L1F_S_ which is heavily dependent on variations in regularization parameter and undersampling factor. The results for the other regularization parameters are included in **Figure S3** in **Supplementary Materials.**

**Figure 5:**
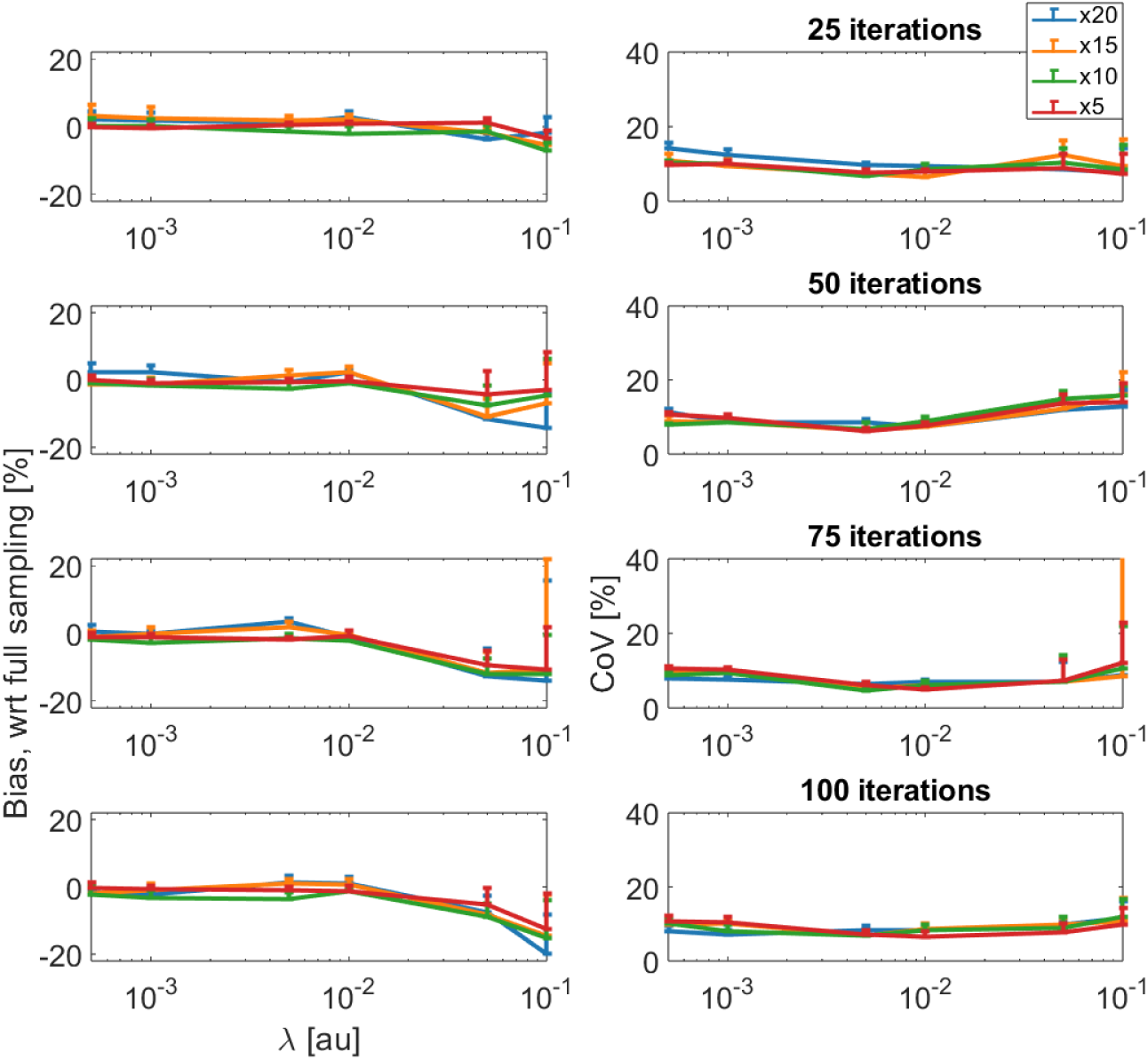
Bias and CoV as a function of regularization parameter across all undersampling factors. This example presents results for T1 maps calculated from images reconstructed using joint L1-wavelet regularization in the spatial and contrast domain. For this regularization operator, uncertainty across T1 maps is minimized at regularization parameters in the range 10^−3^ – 10^−2^ with weak dependence on undersampling factor.

Representative computation times for reconstructions with different number of iterations, regularization parameters, and undersampling factors are shown in **Figure S4** in **Supplementary Materials.**

### 3.3. In Vivo Imaging

A comparison between the benchmark and PICS reconstruction is presented in **Figure 6**. This example uses half-spoke coverage for a total scan time of ∼4.5 minutes. PICS reconstruction time was ∼28 minutes in a high-performance workstation with 8 nodes, each with 18-core Intel Xeon CPUs providing a total of 512 GB of RAM. The respiratory cycle was amplitude-sorted and divided into ten phases. For clarity, **Figure 6a-b** show phases 1, 3, 5, 7, and 9 of the reconstruction. **Figure 6c** plots the respiratory signal extracted from principal component analysis of the amplitude of the k0-signal from all receive channels during the first minute of the acquisition. Respiratory phase sorting based on signal amplitude was used for motion artifact reduction by calculating T1 maps based on the data from respective end-inspiration and end-expiration phases. The image from end-expiration was calculated by pooling k-space data within one respiratory phase from the minimum of the respiratory signal. A similar approach was followed for end-inspiration and k-space data from the maximum of the respiratory signal. Finally, the respective images from the acquisition with each flip angle were used in the calculation of the T1 map.

**Figure 6:**
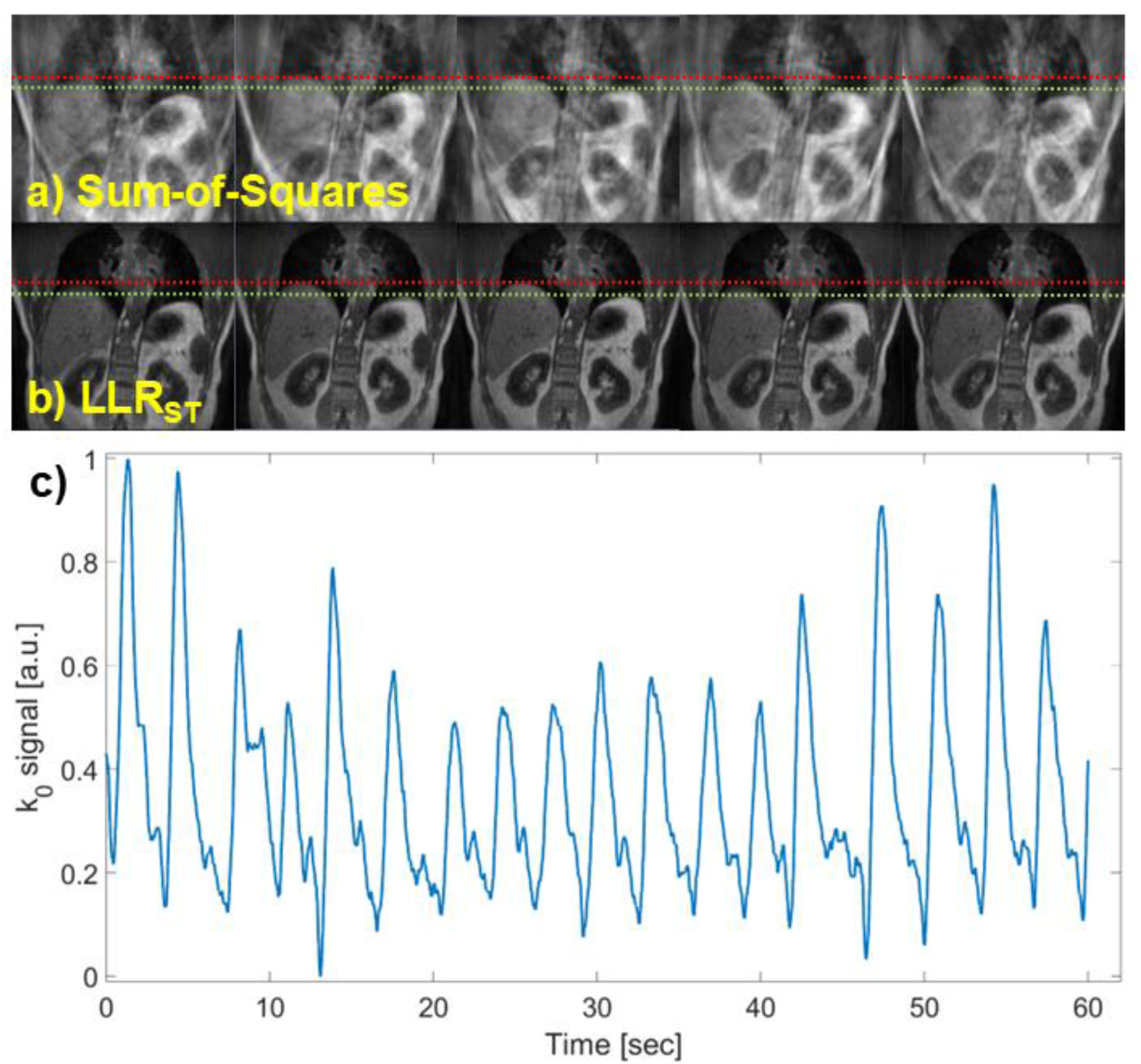
Comparison between the (a) benchmark and (b) PICS reconstruction with LLR_ST_. For clarity, only phases 1, 3, 5, 7, and 9 of the respiratory cycle are shown. The dashed red and green lines show the apex of the liver at end expiration and inspiration, respectively. (c) Respiratory signal extracted from principal component analysis of the amplitude of the k0-signal from all receive channels during the first minute of the acquisition.

**Figure 7** shows a comparison of T1 maps calculated from data acquired with Cartesian sampling and 3D-radial PICS at end-inspiration/expiration using the method outlined above. Mean T1 values at various regions of interest in the volunteer are listed in **Table 4**, which also includes reference T1 values^26^. In the paravertebral muscle, kidney cortex, and kidney medulla mean T1 values exhibit a large bias and uncertainty in the Cartesian acquisition due to strong influence from motion artifacts. Across all k-space sampling methods, the T1 values in the paravertebral muscle are similar since this anatomical region experiences minimal artifacts from respiratory motion. However, in the kidney cortex and kidney medulla, the Cartesian k-space sampling method overestimates the reference values and shows high standard deviation compared to the other k-space sampling methods. In these regions, all non-Cartesian k-space sampling strategies closely follow the reference values. Note the increase in standard deviation in the kidney medulla where the T1 values are higher, reflecting the uncertainty that is also observed in the phantom measurements.

**Figure 7:**
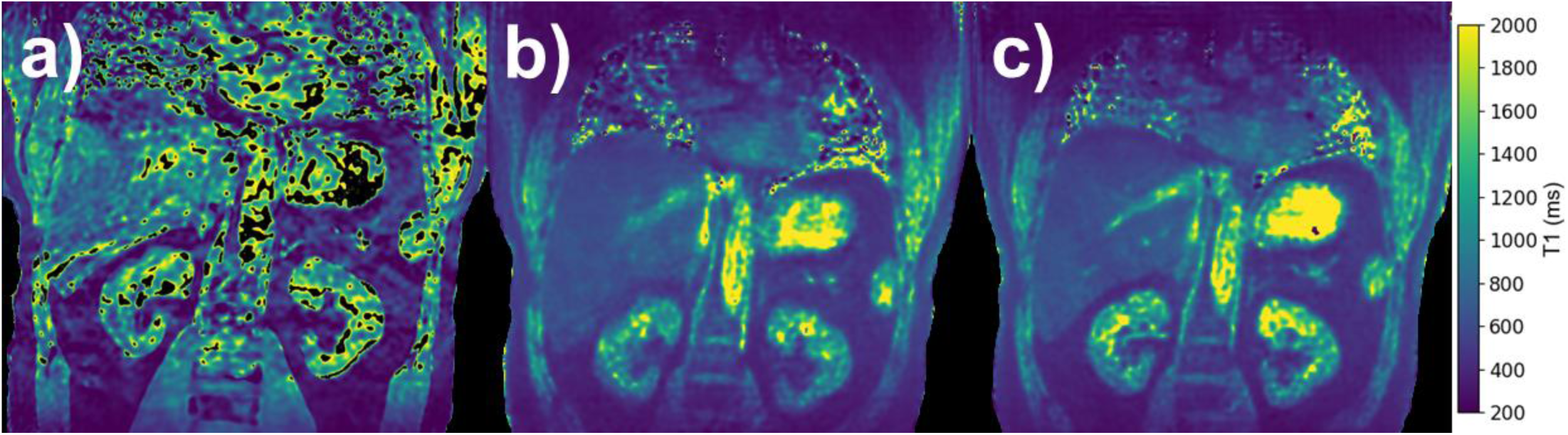
Comparison of T1 maps calculated from data acquired with (a) Cartesian sampling (b) 3D-radial PICS at end-inspiration and (c) 3D-radial PICS at end-expiration. Mean T1 values at various regions of interest in the volunteer are listed in Table 4.

**Table 4:** Comparison of mean and standard deviation of T1 values in stationary and mobile regions. Cartesian sampling overestimates the reference values and shows high standard deviation compared to the non-Cartesian sampling methods in mobile regions due to artifacts from respiratory motion.

|  | Paravertebral Muscle (ms) | Kidney Cortex (ms) | Kidney Medulla (ms) |
| --- | --- | --- | --- |
| **Radial, Half-Spoke** | 829 ± 56 | 971 ± 180 | 1402 ± 361 |
| **Radial, Full-Spoke** | 869 ± 97 | 986 ± 195 | 1494 ± 605 |
| **Stack-of-Stars, Half-Spoke** | 799 ± 50 | 970 ± 219 | 1492 ± 279 |
| **Stack-of-Stars, Full-Spoke** | 949 ± 109 | 999 ± 185 | 1523 ± 199 |
| **Cartesian** | **938 ± 42** | **1383 ± 416** | **2484 ± 2376** |
| **Reference** | **856 ± 61** | **966 ± 58** | **1412 ± 58** |

An implementation of respiratory phase resolved T1-mapping using 3D-radial PICS in a patient undergoing radiation therapy is shown in **Figure 8**. This example uses half-spoke coverage for a total scan time of ∼4.5 minutes. The arrow in **Figure 8a** points to the target in the left kidney. Note the presence of a cyst in the inferior aspect of the contralateral kidney, where the T1 values are mostly outside the range of the T1 that can be estimated with our current method. The T1 values in the kidney cortex, kidney medulla, and tumor were 964 ± 153 ms, 1390 ± 729 ms, and 1073 ± 211 ms, reflecting agreement with reference values.

**Figure 8:**
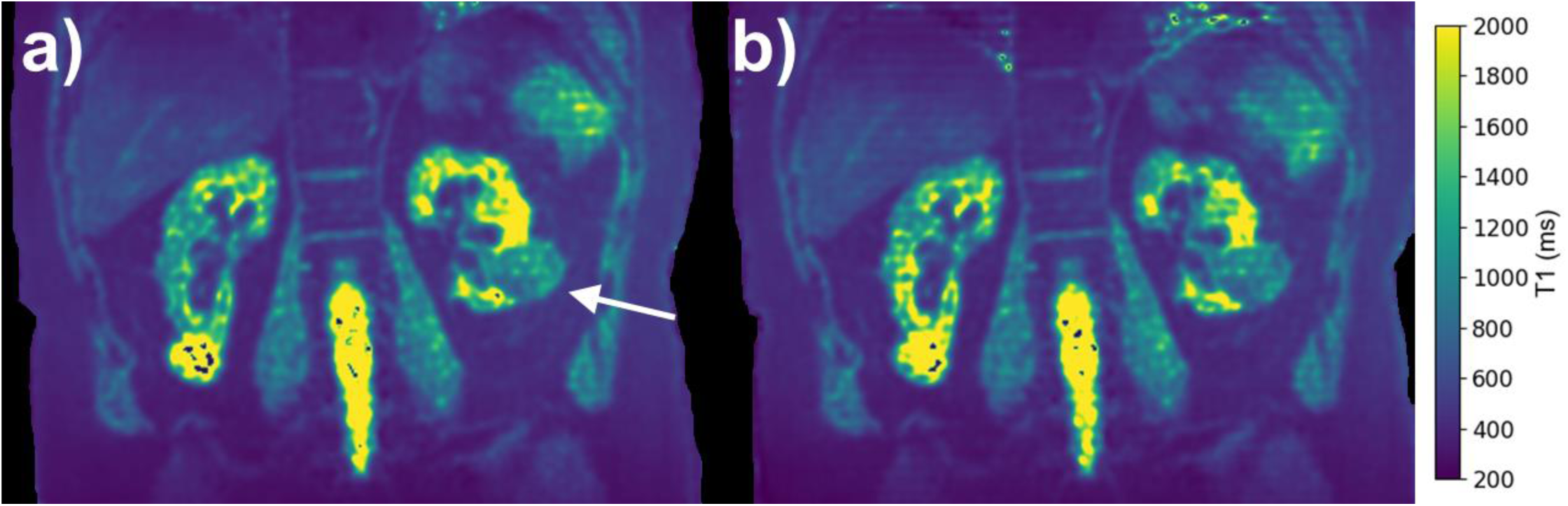
Implementation of respiratory phase resolved T1-mapping using 3D-radial PICS in a patient undergoing radiation therapy to a target in the left kidney, arrow in panel (a). Note the presence of a cyst in the inferior aspect of the contralateral kidney, where the T1 values are mostly outside the range of the T1 that can be estimated with our current method. The T1 values in the kidney cortex, kidney medulla, and tumor were 964 ± 153 ms, 1390 ± 729 ms, and 1073 ± 211 ms, reflecting agreement with reference values.

## 4. Discussion

This study demonstrates the feasibility of implementing robust and clinically compatible longitudinal relaxation rate (R1) mapping on a 1.5T MR-Linac using non-Cartesian sampling strategies and advanced offline compressed sensing (CS) reconstruction. Our results highlight the potential of combining 3D radial acquisitions with tailored regularization schemes to overcome traditional challenges in MR-guided adaptive radiotherapy (MRgART), particularly in anatomical regions subject to large respiratory motion such as the abdomen.

The implementation of T1 mapping on the MR-Linac has traditionally been limited by long scan times, susceptibility to motion artifacts, and a lack of optimized reconstruction techniques. In this work, we addressed these limitations through: (1) the use of radial k-space trajectories that support volumetric and motion-resilient acquisitions; (2) the application of compressed sensing reconstruction with regularization in the spatial and contrast domain; (3) the incorporation of gradient delay correction for minimizing trajectory-mismatch artifacts; and (4) respiratory phase-resolved sorting for retrospective correction of motion-induced artifacts.

Our phantom results confirmed that all tested non-Cartesian methods maintained high reproducibility with minimal longitudinal bias. Among the trajectories evaluated, the half-spoke radial sampling exhibited the best combination of spatial resolution, minimal bias, and shortest acquisition time (∼2.5 min per flip angle). The slightly larger bias observed in the stack-of-stars and full-spoke trajectories at high T1/T2 values is likely due to incomplete spoiling and signal recovery, a known limitation in variable flip angle (VFA) methods using RF-spoiled gradient echo sequences. These observations are consistent with prior reports emphasizing the importance of sequence spoiling in high dynamic range applications, especially when aiming for quantitative fidelity across multiple tissues with diverse relaxation characteristics.

Compressed sensing reconstruction further improved the accuracy and robustness of T1 estimates in undersampled datasets. Notably, regularization strategies that exploit spatiotemporal redundancies—such as joint L1-wavelet transforms (L1W_ST_) and local low-rank methods (LLR_ST_)—were superior in preserving spatial resolution and minimizing parameter-domain uncertainty across all tested undersampling factors. These results support prior work demonstrating the effectiveness of sparsifying transforms and rank-constrained models in enabling dynamic and quantitative MRI with aggressive undersampling. Our findings further validate these techniques within the constraints of the MR-Linac system and for the specific goal of T1 mapping in motion-prone anatomy.

In vivo imaging reinforced the value of non-Cartesian sampling and optimized PICS reconstruction. When compared to Cartesian acquisitions, 3D radial PICS methods provided more accurate and precise T1 estimates in mobile tissues such as the kidney cortex and medulla. This improved performance likely stems from both the intrinsic motion insensitivity of radial sampling and the ability of the CS framework to selectively incorporate motion-resolved data through respiratory sorting. The clinical feasibility of implementing this framework was demonstrated in a patient undergoing radiation therapy, where phase-resolved T1 mapping revealed physiologically relevant contrast and acceptable agreement with reference values. While the current implementation requires substantial offline reconstruction time, further optimization and hardware acceleration (e.g., GPU-enabled pipelines) are anticipated to improve integration into MRgART workflows.

A few limitations should be noted. First, although two phantoms were used to validate the methods, the sample size for in vivo evaluation was limited. Larger studies will be necessary to evaluate inter-subject variability, assess reproducibility across different anatomical sites, and validate longitudinal changes in response to therapy. Second, the accuracy of VFA-based T1 mapping remains sensitive to B1 inhomogeneity and imperfect spoiling, which may necessitate corrections or alternative techniques such as B1 mapping or Look-Locker-based methods in future work. Third, while this study focused on static and respiratory-sorted reconstructions, additional temporal dimensions (e.g., contrast-enhanced or perfusion imaging) could be incorporated using the same framework to enable true multiparametric imaging on the MR-Linac.

Overall, our results support the feasibility and accuracy of non-Cartesian, motion-resolved T1 mapping with compressed sensing on the MR-Linac. This work sets the stage for integrating quantitative MRI into MR-guided adaptive workflows, enabling more biologically informed treatment planning, longitudinal biomarker monitoring, and real-time adaptation based on tissue response.

## 5. Conclusion

This study demonstrates the feasibility of longitudinal relaxation rate (R1) mapping on a 1.5T MR-Linac using clinically available non-Cartesian sampling trajectories combined with advanced compressed sensing reconstruction. Among the evaluated strategies, 3D radial half-spoke sampling coupled with joint spatiotemporal regularization (e.g., L1-wavelet and local low-rank methods) provided the best trade-off between acquisition time, spatial resolution, and quantitative accuracy—supporting robust performance under high undersampling conditions. Gradient delay correction and retrospective respiratory sorting further enhanced image quality and motion robustness.

The proposed methods enable volumetric, motion-resolved T1 mapping in anatomies prone to respiratory motion and were validated across two phantoms, a healthy volunteer, and a clinical case. These capabilities establish a pathway for integrating quantitative MRI into MR-guided adaptive radiotherapy workflows. Future studies will focus on expanding clinical evaluation, optimizing reconstruction speed, and extending the framework to additional quantitative biomarkers and multi-parametric imaging protocols.

## 6. Supplementary Materials

**Figure S1:**
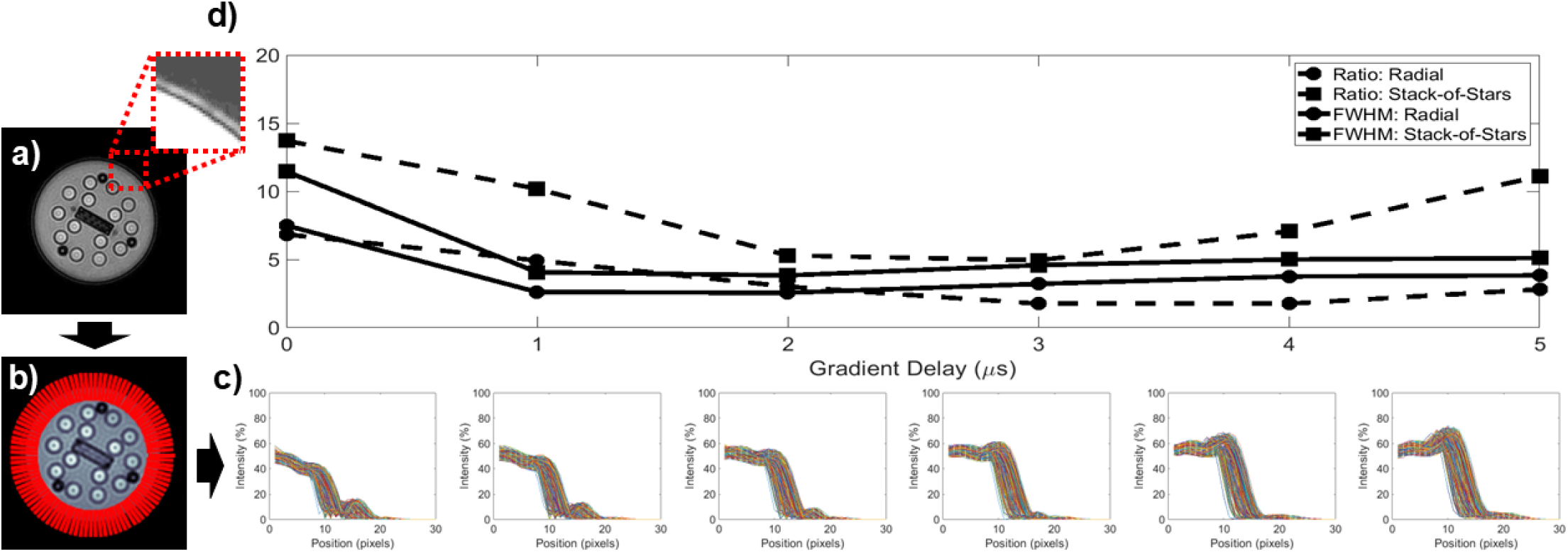
Overview of analysis for gradient delay time optimization. a) Ghosting artifact at the edge of the phantom, b) A set of radial spokes placed every 1° to analyze line profiles, c) Intensity line profiles for each radial spoke, d) Computed ratio and FWHM for both the half-spoke radial and stack-of-stars sequences.

**Figure S2:**
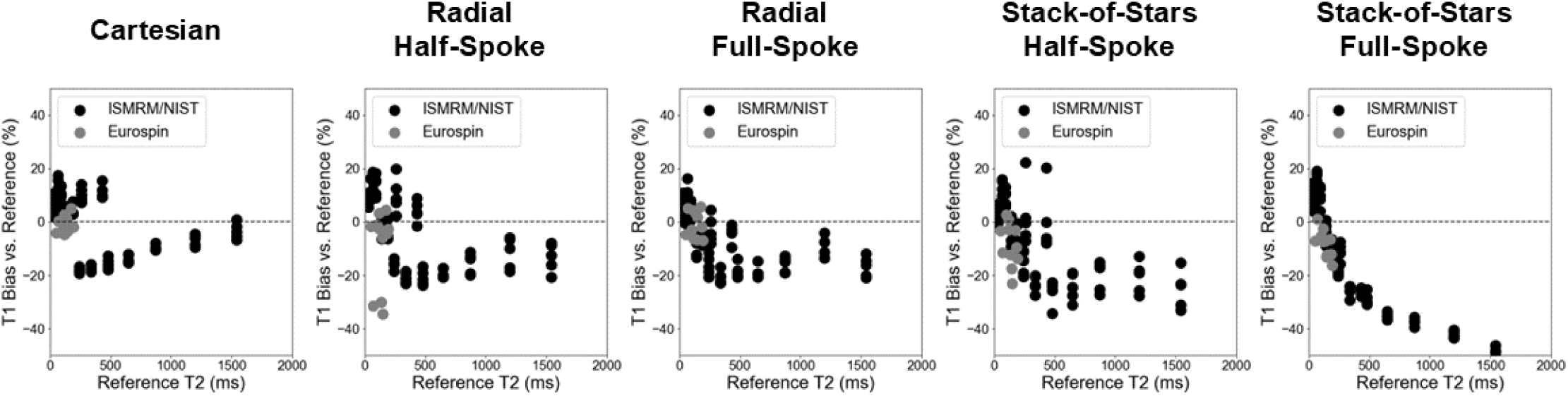
T1 bias as a function of reference T2 values for all vials in the ISMRM/NIST and Eurospin phantom. Note general trend in increase in bias due to insufficient spoiling and signal recovery in vials with high T1 and T2 values.

**Figure S3:**
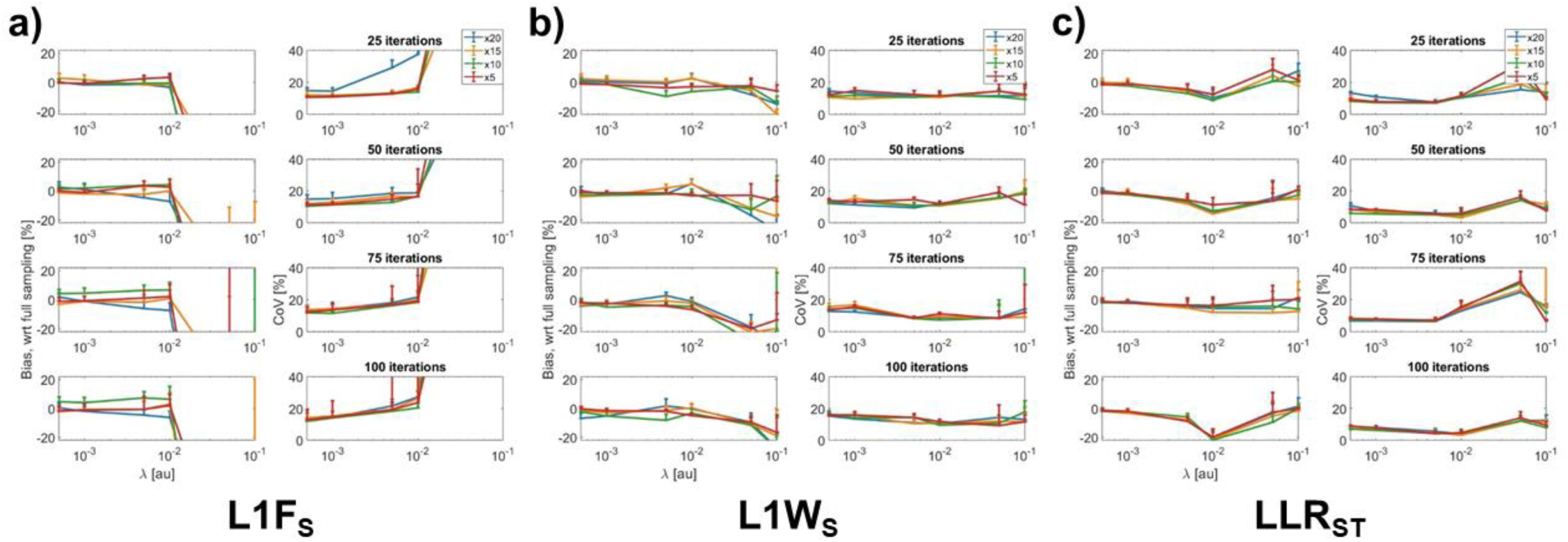
Bias and CoV as a function of regularization parameter across all undersampling factors. The panels present results for T1 maps calculated from images reconstructed using a) L1F_S_, b) L1W_S_, c) LLR_ST_.

**Figure S4:**
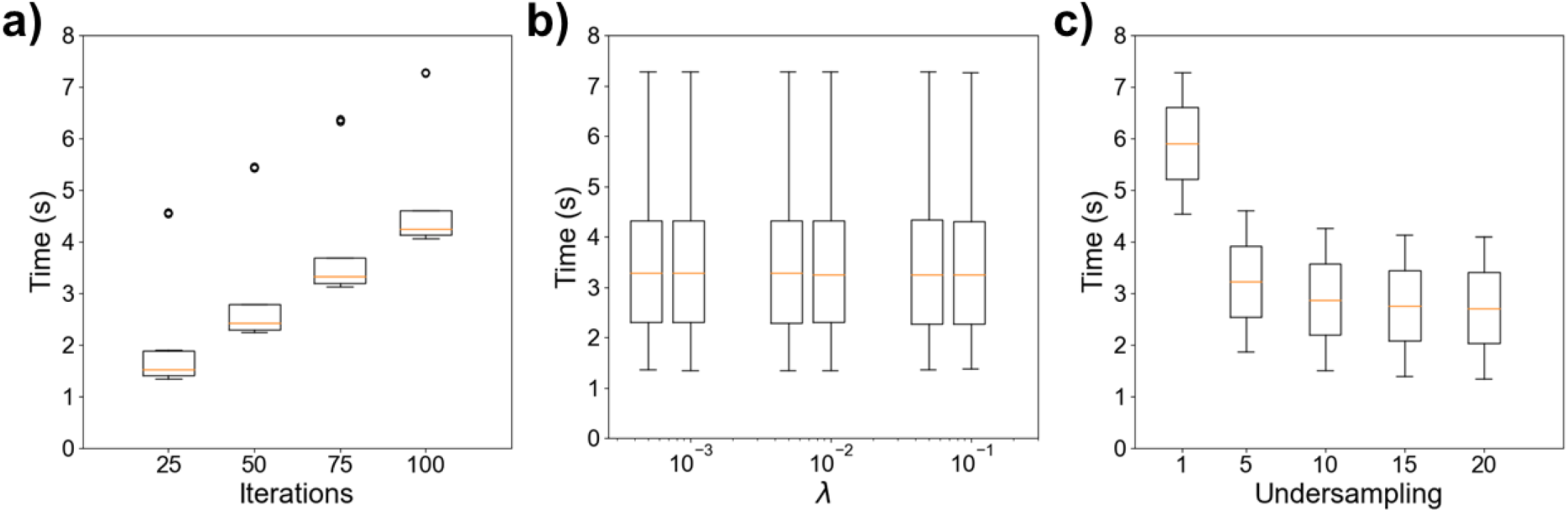
PICS reconstruction time across the number of iterations (a), regularization parameter (b), and undersampling factor (c). Note that the reported values do not include the calculation of the coil sensitivity maps, which in this case was on the order of 8 - 12min. Outlier in panel (a) represents reconstruction for fully sampled dataset.

## Contributor Roles Taxonomy (CRediT) Attribution Statement

*Conceptualization:* L.M. and E.D.S.; *Data curation:* L.M., M.J.V.R., and E.D.S; *Formal analysis:* L.M. and E.D.S.; *Funding acquisition:* E.D.S. and C.D.F.; *Investigation:* L.M., M.J.V.R., and E.D.S.; *Methodology:* L.M. and E.D.S.; *Project administration:* E.D.S.; *Resources:* K.H., Y.D., C.T., C.H., J.Y., P.B., and J.W.; *Software:* L.M. and E.D.S.; *Supervision:* E.D.S. and C.D.F.; *Validation:* L.M., M.J.V.R., and E.D.S.; *Visualization:* L.M. and E.D.S.; *Writing – original draft:* L.M. and E.D.S.; *Writing - review & editing:* L.M., M.J.V.R., K.H., Y.D., C.T., C.H., J.Y., P.B., J.W., C.D.F., and E.D.S.

## Data Availability Statement

All relevant anonymized data necessary for figure reproduction are available at the following FigShare DOI: 10.6084/m9.figshare.29637422. The accompanying code for image visualization and statistical analysis will be made publicly available at the following URL: https://github.com/Lucas-Mc/non-cartesian-relaxometry.

## Acknowledgements

The author(s) acknowledge the support of the High-Performance Computing for research facility at The University of Texas MD Anderson Cancer Center for providing computational resources that have contributed to the research results reported in this paper.

## Author Declarations

All participants provided written informed consent and were consented to an internal imaging protocol (PA15-0418), both approved by the institutional review board at The University of Texas MD Anderson Cancer Center.

